# DNA Methylation Signatures of Atherosclerosis and Vascular-Related Outcomes in U.S. and Irish Population-Based Cohorts

**DOI:** 10.64898/2026.05.25.26354072

**Authors:** Farah Ammous, Trey Smith, Siobhan Scarlett, Belinda Hernández, Cathal McCrory, Rose Anne Kenny, Colter Mitchell, Jessica D. Faul

## Abstract

Atherosclerosis is a systemic vascular process linked to cardiovascular, cognitive and renal outcomes. DNA methylation (DNAm)-based scores of atherosclerosis may capture cumulative biological processes underlying vascular aging. Here, we examined associations of DNAm scores for coronary artery calcification (DNAm-CAC) and carotid plaque (DNAm-cPlaque), derived from a large study of imaging-based subclinical atherosclerosis, with prevalent and incident outcomes in two population-based cohorts of older adults: the Health and Retirement Study (HRS; n = 3,875) and The Irish Longitudinal Study on Ageing (TILDA; n = 487). Higher DNAm scores were associated with adverse cardiometabolic profiles and socioeconomic indicators. In HRS, higher DNAm-CAC was associated with prevalent cardiovascular disease (odds ratio per SD, 1.16; 95% confidence interval (CI), 1.07–1.26), lower cognitive function (β =-0.50, 95% CI-0.68 to-0.32) and lower estimated glomerular filtration rate (eGFR; −1.7 ml min^-1^ 1.73 m^-2^, 95% CI-2.6 to-0.8) in unadjusted models. After adjustment for demographic and clinical risk factors, DNAm-CAC (β =-0.29, 95% CI-0.46 to-0.13) and DNAm-cPlaque (β =-0.24, 95% CI-0.42 to-0.06) remained associated with lower cognitive function, and DNAm-cPlaque was associated with incident cognitive impairment or dementia (hazard ratio per SD, 1.16; 95% CI, 1.01–1.32). Associations were attenuated after further adjustment for race/ethnicity and socioeconomic indicators. In TILDA, higher DNAm-cPlaque was associated with worse cognitive performance (incidence rate ratio, 1.11; 95% CI, 1.01–1.21), increased risk of incident cardiovascular disease (hazard ratio, 1.18; 95% CI, 1.00–1.42) and lower eGFR, with consistent associations observed for DNAm-CAC. These findings suggest that DNAm-based scores of atherosclerosis capture systemic vascular processes linked to multiple age-related outcomes across populations. Further work is needed to clarify the biological pathways reflected by these scores and their relation to cumulative and socially patterned vascular risk.

## INTRODUCTION

Atherosclerosis is one of the leading causes of morbidity and mortality in the United States and globally.^1,2^ It is a progressive systemic inflammatory disease characterized by the accumulation of lipid-rich and fibrotic material within arterial walls, resulting in plaque formation and vascular calcification.^1,3^ Beyond its role in symptomatic cardiovascular disease (CVD), atherosclerosis contributes to dysfunction in other organ systems through impaired vascular perfusion, including the brain and kidneys. Measures of atherosclerosis, such as coronary artery calcium (CAC), assessed non-invasively using computed tomography, have demonstrated clinical utility for identifying individuals at elevated risk of cardiovascular events and guiding preventive therapies.^4–6^ Atherosclerosis measures have also been associated with poorer cognitive performance, increased risk of dementia, and reduced hippocampal volume.^7–9^ In addition, population-based studies have reported graded associations between CAC burden and reduced kidney function, albuminuria, and chronic kidney disease progression.^10–12^ These findings support the concept that renal impairment may represent a manifestation of systemic vascular injury involving shared mechanisms such as endothelial dysfunction, arterial stiffness, vascular calcification, and chronic inflammation.

Despite advances in imaging-based assessment, direct measurement of atherosclerosis remains challenging in large population studies due to cost, feasibility, and limited availability of repeated measures. Epigenetic biomarkers, particularly DNA methylation (DNAm), have emerged as promising tools for capturing biological responses to cumulative environmental, behavioral, and social exposures. DNAm is a tissue-specific regulatory mechanism that reflects both genetic influences and long-term exposure to physiological and psychosocial stressors.^13–15^ Increasingly, DNAm-based composite measures have been developed to represent biological aging processes and disease-related risk profiles.^16–19^ Through their sensitivity to social and environmental contexts, epigenetic signatures may provide insight into pathways linking socioeconomic disadvantage with disparities in vascular disease burden.^20–22^

In prior work in a community-based sample of African American participants, we found that epigenetic scores derived from an epigenome-wide association study (EWAS) of atherosclerosis conducted in the Multi-Ethnic Study of Atherosclerosis (MESA) were associated with imaging-based measures of atherosclerosis across multiple vascular beds beyond traditional risk factors.^23^ However, it remains unclear whether such DNAm signatures of atherosclerosis are associated with clinically relevant vascular outcomes across diverse populations and organ systems. Here, we conceptualized DNAm atherosclerosis scores as integrative molecular indicators of systemic vascular aging with potential relevance across multiple vascular-related outcomes. We evaluated the same EWAS-derived scores for coronary artery calcification (DNAm-CAC) and carotid plaque (DNAm-cPlaque) in relation to cardiovascular disease, cognitive function, and kidney function in two population-based cohorts of older adults from the Health and Retirement Study (HRS) and The Irish Longitudinal Study on Ageing (TILDA). We additionally examined the scores’ associations with incident cardiovascular disease and cognitive impairment in the HRS and incident cardiovascular disease in TILDA.

## METHODS

### Study participants

The HRS is a national longitudinal panel study that surveys a representative sample of approximately 20 000 U.S. adults over age 50 every two years.^24^ The HRS is sponsored by the National Institute on Aging (grant number NIA U01AG009740) and is conducted by the University of Michigan. The Venous Blood Study (VBS) was completed in 2016 in which venous blood draws were obtained for biomarker assessment (N = 9934) and for DNA methylation measurement in a sub-sample of participants.^25^ In this analysis, we limited the sample to age-eligible participants with non-zero sampling weights from the 2016 VBS, representing U.S. adults aged 56 years and older (N = 3,875). This analysis included four waves of HRS survey data from 2014 to 2020. All participants provided informed consent to participate in the study. The University of Michigan Institutional Review Board approved the collection and analyses of these data (HUM00061128, HUM00056464, and HUM00192059).

TILDA is a nationally representative cohort of 8175 community-dwelling adults aged ≥50 years which was established in 2006 with the goal of capturing the social, health, and economic circumstances of the aging population in Ireland.^26,27^ The study has been harmonized with leading international research, including the HRS, to ensure comparability of results. Since baseline (Wave 1; 2009–2011), participants have been followed at regular intervals through computer-assisted personal interviews and nurse-led health assessments. TILDA is conducted by Trinity College Dublin and supported by Irish government funding agencies and other sources. At Wave 3 (2014-2015), venous blood samples were obtained for biomarker assessment and DNAm measurement in a subsample of participants (N=487). The DNAm sample was selected based on respondents’ life-course social class trajectory derived from the cross-classification of fathers’ and respondents’ own social class. In the present analysis, we limited the sample to participants with available DNAm data and included longitudinal survey data from Wave 3 through follow-up waves conducted up to 2021. Additional cohort details and selection of the epigenetic sample are described in detail elsewhere.^28,29^ All participants provided written informed consent. Ethical approval for TILDA was granted by the Faculty of Health Sciences Research Ethics Committee in Trinity College Dublin.

### Outcome Assessment

In the HRS, prevalent CVD was defined based on participants’ self-report of a physician diagnosis of heart disease through the 2016 wave. Participants were asked whether a doctor had ever told them they had experienced a heart attack, coronary heart disease, angina, congestive heart failure, other heart problems, or stroke. Kidney function was assessed using estimated glomerular filtration rate (eGFR) calculated from serum cystatin C measured in venous blood samples collected as part of the 2016 VBS using a particle-enhanced immunonephelometric assay on a Roche COBAS 6000 chemistry analyzer.^30^ Cognitive function was assessed using the Langa-Weir classification for self-and proxy-respondents which is publicly available.^31–33^ Participants were assigned a 27-point total summary score for cognition using measures from the core HRS interviews. We categorized the score into three categories: normal cognitive function (score: 12-27), cognitive impairment but not dementia (CIND) (score: 7-11), and dementia (score: 0-6). We also used cognitive status reported by informants for proxy respondents to reduce bias that would otherwise be present from sample attrition. Proxy scores are based on an 11-point score scale: normal (score: 0-2), CIND (score: 3-5), and dementia (6-11). To account for mode differences, scores from the 2018 and 2020 core survey were adjusted for web mode by subtracting 1 from the total cognition score as necessary.^33^

In TILDA, prevalent CVD was defined based on self-reported lifetime physician diagnosis of angina, myocardial infarction, heart failure, stroke, transient ischemic attack, or heart murmur. Cognitive functioning was assessed using the Montreal Cognitive Assessment (MoCA) tool. Cognitive performance was modeled using the number of errors (30 minus the total score), with higher values indicating poorer cognitive function. Kidney function was assessed using eGFR calculated from serum cystatin C measured from non-fasting venous blood samples collected during the health assessment and processed in a central laboratory using standardized clinical chemistry assays.

For incident outcomes, participants with prevalent disease at baseline were excluded. In HRS, incident CVD and incident cognitive impairment or dementia were identified during follow-up waves conducted in 2018 and 2020 based on the Langa-Weir classification as previously described. In TILDA, incident CVD events were ascertained during subsequent waves using comparable self-reported physician diagnoses up to 2021.

### DNA Methylation and Epigenetic Scores Construction

In both the HRS and TILDA, DNAm was profiled in whole blood using the Illumina Infinium MethylationEPIC array. For this analysis, we focused on 82 DNAm sites, known as cytosine-phosphate-guanine (CpG) dinucleotides, previously reported to be associated with CAC or carotid plaque in an epigenome-wide association analysis of 1,208 participants from the Multi-Ethnic Study of Atherosclerosis (MESA) at a false discovery rate < 0.1.^34^ Of the reported CpGs, 75 were available in the EPIC V1.0 array. Missing CpG values were imputed to the sample median for each cohort. Linear mixed models were used to adjust for batch as random effects. We constructed the epigenetic scores as the sum of the methylation levels at each CpG weighted by the EWAS reported effect sizes ^34^. DNAm-CAC was based on 15 CpGs and DNAm-cPlaque was based on 62 CpGs. Scores were standardized to a standard normal distribution, and for each score, outliers beyond six standard deviations (SD) from the mean were excluded (n=2 for DNAm-CAC and n=2 for DNAm-cPlaque in the HRS). The CpGs included in each of the scores are listed in **Table S1** in the **Supplement**.

Additional details on laboratory processing, data preprocessing, quality control, estimation of leukocyte cell proportions, and covariate assessment (including demographic, behavioral, and clinical factors) are described in the **Supplemental Methods.**

### Statistical analysis

In the HRS, cross-sectional associations between DNAm atherosclerosis scores and prevalent CVD were examined using weighted logistic regression models to account for the DNA methylation subsampling design. DNAm-CAC and DNAm-cPlaque scores were modeled as continuous predictors per 1-SD higher score. Sequential adjustment models were fitted to evaluate attenuation of associations. Model 1 included the DNAm score only. Model 2 additionally adjusted for age, sex, smoking status, body mass index (BMI), systolic blood pressure (SBP), diastolic blood pressure (DBP), diabetes, and total cholesterol. Model 3 further adjusted for race/ethnicity and socioeconomic indicators (educational attainment and total household wealth). Model 4 additionally adjusted for estimated leukocyte composition (CD8 naïve T cells, natural killer cells, monocytes, B cells, and CD8 T cell), to assess potential confounding by variation in blood cell proportions. Continuous outcomes, including cognitive function score and eGFR, were analyzed using weighted linear regression models with the same sequential adjustment blocks. Cognitive models additionally adjusted for *APOE* ε4 carrier status after excluding participants with the ε2/ε4 genotype. For incident outcomes, participants with prevalent disease at baseline (2016) were excluded. Time-to-event was defined from the 2016 interview date to the date of event occurrence, death, or last follow-up interview (2018 or 2020), whichever occurred first. Weighted Cox proportional hazards models were used to estimate hazard ratios (HRs) and 95% confidence intervals. The proportional hazards assumption was evaluated using Schoenfeld residuals.

In TILDA, cross-sectional associations with prevalent CVD were evaluated using logistic regression models. MoCA errors were modeled using negative binomial regression due to overdispersion and eGFR was analyzed using linear regression. Incident CVD was analyzed using Weibull accelerated failure time (AFT) models with left, interval, and right censoring. Specifically, participants with CVD at baseline were left-censored; incident cases occurring between study waves were interval-censored; and those without CVD were right-censored at the last observed time or death, whichever occurred first. Hazard ratios were derived from the AFT coefficients as exp(−𝛽/𝜎), where 𝜎 is the model scale parameter. Adjustment models paralleled those used in HRS, with covariates included as available. Model 1 included the DNAm score only. Model 2 additionally adjusted for age, sex, smoking status, BMI, SBP, DBP, diabetes, and total cholesterol. Model 3 further adjusted for educational attainment and household wealth. Model 4 additionally adjusted for estimated leukocyte proportions (granulocytes, CD4 T cells, CD8 T cells, B cells, monocytes, and natural killer cells). A sensitivity analysis using Cox proportional hazards models was conducted to evaluate the robustness of the AFT model results. Sample sizes varied across models due to outcome-specific eligibility and covariate availability.

All statistical analyses were conducted separately in the HRS and TILDA using R. Weighted analyses in HRS were performed using the *survey* package.^35^ Effect estimates are presented per 1-SD higher DNAm score with 95% confidence intervals. Statistical significance was defined using two-sided *P* values < 0.05.

## RESULTS

### Participant Characteristics

The analytical sample included 3,875 participants from the HRS and 487 participants from TILDA **(Table 1)**. Compared with HRS, the TILDA sample was younger and included a lower proportion of women. The HRS sample was diverse with weighted percentages of approximately 78% non-Hispanic White, followed by 10% non-Hispanic Black and 9% Hispanic **(Table S2).** In TILDA, information on participants’ race/ethnicity were not available; however, the cohort is predominantly composed of individuals of White European background. The TILDA sample had lower BMI, higher average blood pressure, and lower prevalence of diabetes. The outcome burden was also lower in TILDA, with lower prevalent CVD (14.3% vs 32.2%) and higher eGFR (80.6 vs 64.8).

**Table 1.**
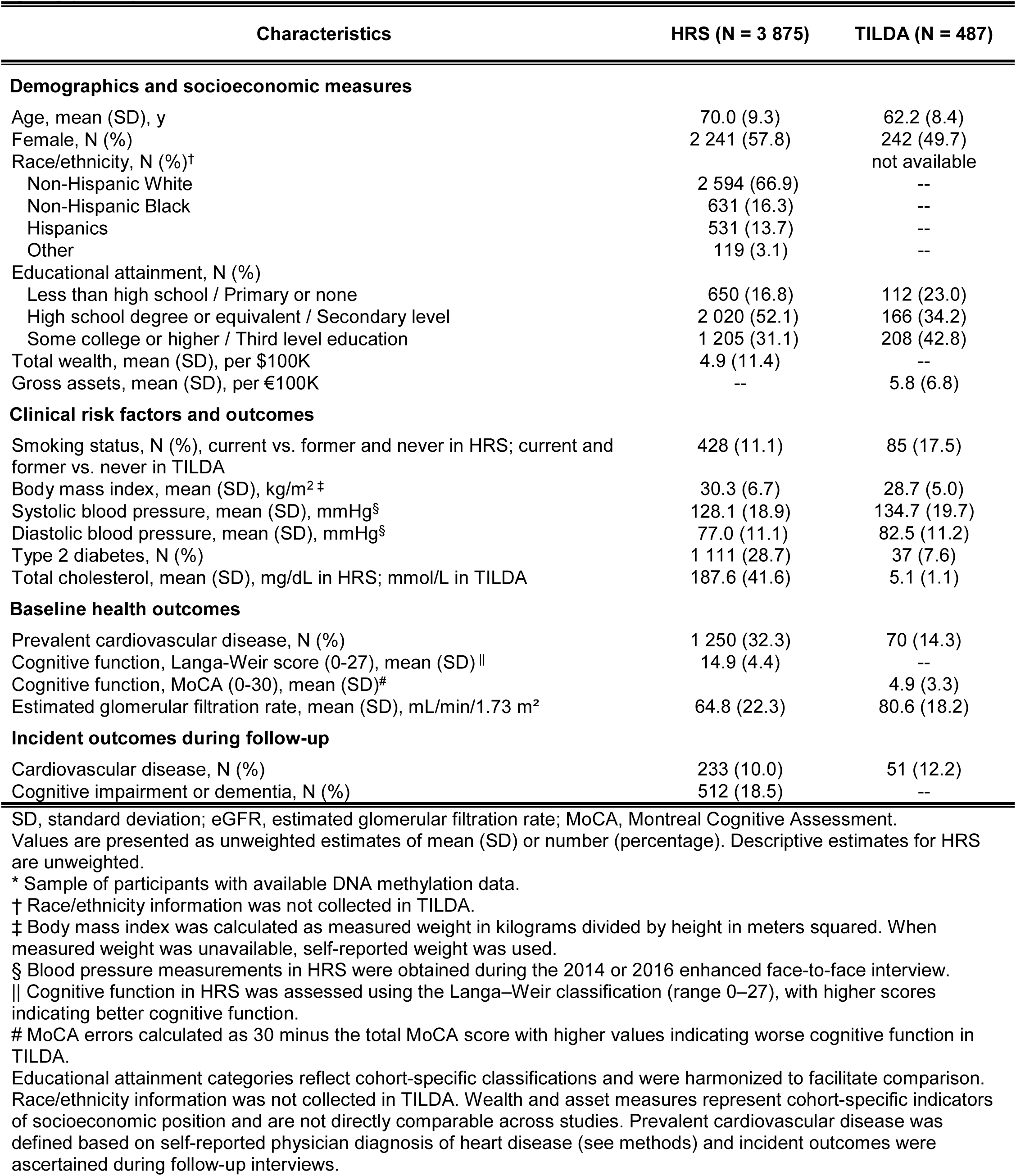
Participant Characteristics in the Health and Retirement Study (HRS) and The Irish Longitudinal Study on Ageing (TILDA) *

| Characteristics | HRS (N = 3 875) | TILDA (N = 487) |
| --- | --- | --- |
| **Demographics and socioeconomic measures** |  |  |
| Age, mean (SD), y | 70.0 (9.3) | 62.2 (8.4) |
| Female, N (%) | 2 241 (57.8) | 242 (49.7) |
| Race/ethnicity, N (%)† |  | not available |
| Non-Hispanic White | 2 594 (66.9) | -- |
| Non-Hispanic Black | 631 (16.3) | -- |
| Hispanics | 531 (13.7) | -- |
| Other | 119 (3.1) | -- |
| Educational attainment, N (%) |  |  |
| Less than high school / Primary or none | 650 (16.8) | 112 (23.0) |
| High school degree or equivalent / Secondary level | 2 020 (52.1) | 166 (34.2) |
| Some college or higher / Third level education | 1 205 (31.1) | 208 (42.8) |
| Total wealth, mean (SD), per \$100K | 4.9 (11.4) | -- |
| Gross assets, mean (SD), per €100K | -- | 5.8 (6.8) |
| **Clinical risk factors and outcomes** |  |  |
| Smoking status, N (%), current vs. former and never in HRS; current and former vs. never in TILDA | 428 (11.1) | 85 (17.5) |
| Body mass index, mean (SD), kg/m<sup>2</sup>‡ | 30.3 (6.7) | 28.7 (5.0) |
| Systolic blood pressure, mean (SD), mmHg§ | 128.1 (18.9) | 134.7 (19.7) |
| Diastolic blood pressure, mean (SD), mmHg§ | 77.0 (11.1) | 82.5 (11.2) |
| Type 2 diabetes, N (%) | 1 111 (28.7) | 37 (7.6) |
| Total cholesterol, mean (SD), mg/dL in HRS; mmol/L in TILDA | 187.6 (41.6) | 5.1 (1.1) |
| **Baseline health outcomes** |  |  |
| Prevalent cardiovascular disease, N (%) | 1 250 (32.3) | 70 (14.3) |
| Cognitive function, Langa-Weir score (0-27), mean (SD) \|\| | 14.9 (4.4) | -- |
| Cognitive function, MoCA (0-30), mean (SD) # |  | 4.9 (3.3) |
| Estimated glomerular filtration rate, mean (SD), mL/min/1.73 m<sup>2</sup> | 64.8 (22.3) | 80.6 (18.2) |
| **Incident outcomes during follow-up** |  |  |
| Cardiovascular disease, N (%) | 233 (10.0) | 51 (12.2) |
| Cognitive impairment or dementia, N (%) | 512 (18.5) | -- |

SD, standard deviation; eGFR, estimated glomerular filtration rate; MoCA, Montreal Cognitive Assessment.
Values are presented as unweighted estimates of mean (SD) or number (percentage). Descriptive estimates for HRS are unweighted.
\* Sample of participants with available DNA methylation data.
† Race/ethnicity information was not collected in TILDA.
‡ Body mass index was calculated as measured weight in kilograms divided by height in meters squared. When measured weight was unavailable, self-reported weight was used.
§ Blood pressure measurements in HRS were obtained during the 2014 or 2016 enhanced face-to-face interview.
|| Cognitive function in HRS was assessed using the Langa-Weir classification (range 0–27), with higher scores indicating better cognitive function.

### Determinants of DNAm Atherosclerosis Scores

Associations between DNAm atherosclerosis scores and sample characteristics are shown in **Table 2** for the HRS and **Table 3** for TILDA. DNAm-CAC was positively associated with age in both cohorts. In HRS, both DNAm atherosclerosis scores were socially patterned, varying across race/ethnicity, educational attainment, and household wealth. For example, mean DNAm-CAC scores were higher among participants with lower educational attainment and lower wealth. In TILDA, an educational gradient was also observed, with higher DNAm atherosclerosis scores among participants with lower educational attainment. In both cohorts, scores were higher among current and former smokers compared with never smokers. Associations with BMI differed across cohorts: higher BMI was associated with lower DNAm-cPlaque in HRS, whereas higher BMI was associated with higher DNAm-CAC in TILDA. In the HRS, at least one DNAm score was associated with blood pressure, diabetes, and cholesterol measures, whereas similar associations were not observed in TILDA.

**Table 2.** Associations of participant characteristics with DNAm atherosclerosis scores in the Health and Retirement Study (HRS)

|  | DNAm-CAC |  | DNAm-cPlaque |  |
| --- | --- | --- | --- | --- |
|  | Beta (SE) | P value | Beta (SE) | P value |
| Age | 0.008 (0.002) | $2.2 \times 10^{-5}$ | 0.001 (0.002) | 0.52 |
| Female gender | 0.016 (0.037) | 0.67 | 0.0004 (0.039) | 0.99 |
| Race/ethnicity |  |  |  |  |
| Non-Hispanic Whites | Ref. |  | Ref. |  |
| Non-Hispanic Blacks | 0.54 (0.06) | $2.3 \times 10^{-20}$ | 0.41 (0.06) | $1.5 \times 10^{-10}$ |
| Hispanics | 0.17 (0.06) | $4.9 \times 10^{-3}$ | 0.13 (0.06) | 0.02 |
| Other | 0.19 (0.10) | $5.0 \times 10^{-3}$ | 0.093 (0.07) | 0.77 |
| Educational attainment |  |  |  |  |
| Less than high school | Ref. |  | Ref. |  |
| High school degree or equivalent | -0.25 (0.06) | $7.2 \times 10^{-6}$ | -0.22 (0.06) | $9.5 \times 10^{-5}$ |
| Some college or higher | -0.35 (0.06) | $3.1 \times 10^{-9}$ | -0.41 (0.06) | $3.9 \times 10^{-12}$ |
| Total wealth, per \$100K | -0.004 (0.001) | $3.0 \times 10^{-4}$ | -0.005 (0.002) | $5.0 \times 10^{-3}$ |
| Current smoker (vs. never/former) | 0.19 (0.062) | 0.002 | 1.09 (0.055) | $1.9 \times 10^{-82}$ |
| Body mass index, kg/m<sup>2</sup> | 0.002 (0.003) | 0.56 | -0.012 (0.003) | $1.1 \times 10^{-5}$ |
| Systolic blood pressure, mmHg | 0.002 (0.001) | 0.09 | 0.003 (0.001) | $9.0 \times 10^{-3}$ |
| Diastolic blood pressure, mmHg | 0 (0.002) | 0.99 | 0.005 (0.002) | $2.0 \times 10^{-3}$ |
| Type 2 diabetes | 0.16 (0.040) | $6.8 \times 10^{-5}$ | 0.05 (0.042) | 0.40 |
| Total cholesterol, mg/dL | -0.001 (0.0004) | $9.0 \times 10^{-3}$ | 0.0004 (0.001) | 0.40 |

Effect estimates represent the change in DNAm score per one-unit increase in continuous variables or relative to the reference category for categorical variables.
Models were estimated using survey weights to account for the complex sampling design of HRS.
Models were estimated using complete-case analysis among participants with available DNA methylation and covariate data.

**Table 3.** Associations of participant characteristics with DNAm atherosclerosis scores in The Irish Longitudinal Study on Ageing (TILDA)

|  | DNAm-CAC |  | DNAm-cPlaque |  |
| --- | --- | --- | --- | --- |
|  | Beta (SE) | P value | Beta (SE) | P value |
| Age | 0.018 (0.005) | $1.0 \times 10^{-3}$ | 0.006 (0.005) | 0.26 |
| Female gender | -0.032 (0.091) | 0.73 | 0.089 (0.091) | 0.33 |
| Educational attainment |  |  |  |  |
| primary/none | Ref. | -- | Ref. | -- |
| Secondary | -0.191 (0.122) | 0.12 | -0.221 (0.121) | 0.07 |
| Third level | -0.283 (0.117) | 0.02 | -0.353 (0.116) | $3.0 \times 10^{-3}$ |
| Total wealth | -0.003 (0.007) | 0.65 | -0.011 (0.007) | 0.10 |
| Current and former smoker (vs. never) | 0.288 (0.119) | 0.02 | 1.259 (0.105) | $3.2 \times 10^{-29}$ |
| Body mass index (kg/m<sup>2</sup>) | 0.022 (0.009) | 0.01 | 0.004 (0.009) | 0.70 |
| Systolic blood pressure (mmHg) | 0.002 (0.002) | 0.33 | 0.002 (0.002) | 0.42 |
| Diastolic blood pressure (mmHg) | 0.005 (0.004) | 0.22 | 0.002 (0.004) | 0.55 |
| Type 2 diabetes | 0.275 (0.178) | 0.12 | 0.153 (0.178) | 0.39 |
| Total cholesterol (mg/dL) | -0.008 (0.041) | 0.85 | 0.077 (0.041) | 0.06 |

Effect estimates represent the change in DNAm score per one-unit increase in continuous variables or relative to the reference category for categorical variables
Models were estimated using complete-case analysis among participants with available DNA methylation and covariate data.

### Associations of DNAm Atherosclerosis Scores with Cross-Sectional Outcomes

Cross-sectional associations of DNAm atherosclerosis scores with vascular-related outcomes are shown in **Figure 1**. In the HRS, higher DNAm-CAC was associated with greater vascular disease burden, including higher odds of prevalent CVD, lower cognitive function, and lower eGFR. Each SD increase in DNAm-CAC was associated with 17% higher odds of CVD, a 0.5-point lower Langa–Weir cognitive score, and a 1.7 mL/min/1.73 m² lower eGFR in unadjusted models. Adjustment for demographic, clinical, lifestyle, and socioeconomic factors progressively attenuated these associations, with the greatest attenuation observed after adjustment for socioeconomic characteristics (Model 3, **Table S2**). DNAm-cPlaque showed weaker and more selective associations. Higher DNAm-cPlaque was associated with lower cognitive performance in minimally adjusted models but was not clearly associated with prevalent CVD or eGFR. Similar to DNAm-CAC, effect estimates for cognitive function were attenuated after covariate adjustment (Model 3) and were not evident in fully adjusted models.

**Figure 1.**
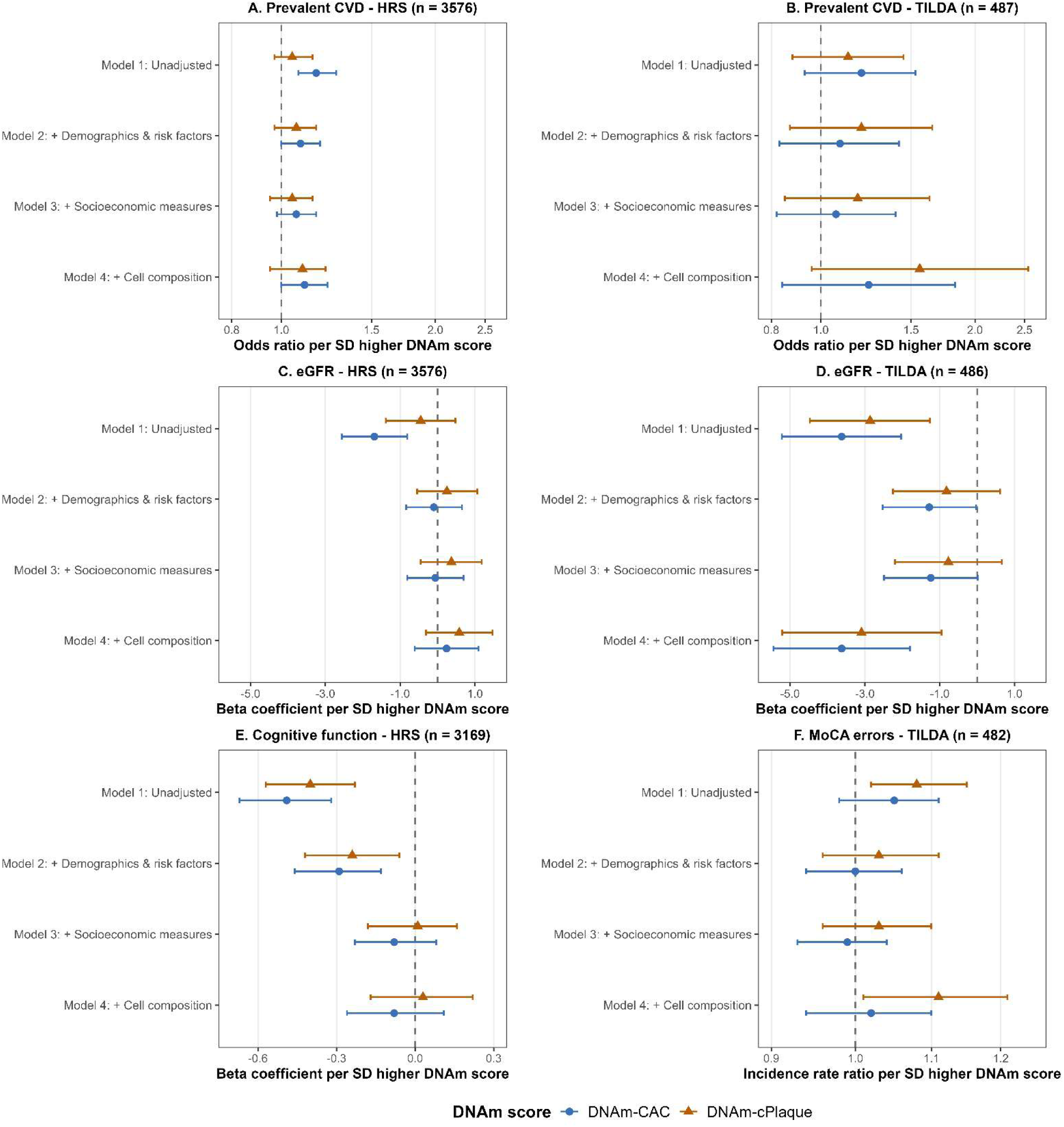
Associations of DNAm atherosclerosis scores with cross-sectional vascular outcomes in the HRS and TILDA. Panels. **A** and **B** show prevalent cardiovascular disease (CVD); **Panels C** and **D**, estimated glomerular filtration rate (eGFR); and **Panels E** and **F** show cognitive outcomes, modeled as Langa-Weir cognitive function score in HRS and MoCA errors in TILDA. Effect estimates and 95% CIs are shown for DNAm-CAC and DNAm-cPlaque across sequential adjustment models. Model 1 was unadjusted. Model 2 adjusted for age, sex, and clinical/lifestyle risk factors. Model 3 additionally adjusted for socioeconomic measures. Model 4 additionally adjusted for estimated blood cell composition. Cognitive estimates are β coefficients for the Langa-Weir score in HRS and incidence rate ratios for MoCA errors in TILDA (corresponding estimates are provided in **Table S3** and **Table S4**).

In TILDA, neither DNAm score showed clear associations with prevalent CVD across adjustment models, although point estimates suggested higher odds of CVD with higher DNAm scores. DNAm-cPlaque was associated with poorer cognitive performance in minimally adjusted models and after adjustment for estimated leukocyte proportions, but not in models with only clinical risk factors and educational attainment. Specifically, each SD increase in DNAm-cPlaque was associated with 8-11% more MoCA errors. The most consistent cross-sectional signal in TILDA was observed for kidney function, where higher DNAm-CAC and DNAm-cPlaque were both associated with lower eGFR and remained significant in fully adjusted models, with each SD increase associated with 3.6 and 3.1 mL/min/1.73m^2^ lower eGFR, respectively.

### Associations of DNAm Atherosclerosis Scores with Incident Outcomes

We next evaluated prospective associations between DNAm atherosclerosis scores and incident vascular-related outcomes **(Figure 2)**. In the HRS, 2329 participants were free of CVD at baseline and 233 incident CVD events occurred over a mean follow-up of 3.6 years (SD=0.8). DNAm scores were not significantly associated with incident CVD across adjustment models, although effect estimates were generally in the expected direction. The analytical sample for cognitive impairment outcomes included 2837 participants, among whom 512 developed CIND or dementia over a mean follow-up of 3.4 years (SD=1.2). Both higher DNAm-CAC and DNAm-cPlaque were associated with approximately 18% and 24% higher risk of incident cognitive impairment or dementia in the minimally adjusted models, respectively. Consistent with cross-sectional findings, these associations progressively attenuated after adjustment for clinical risk factors, with the greatest reduction observed after accounting for socioeconomic characteristics (Model 3).

**Figure 2.**
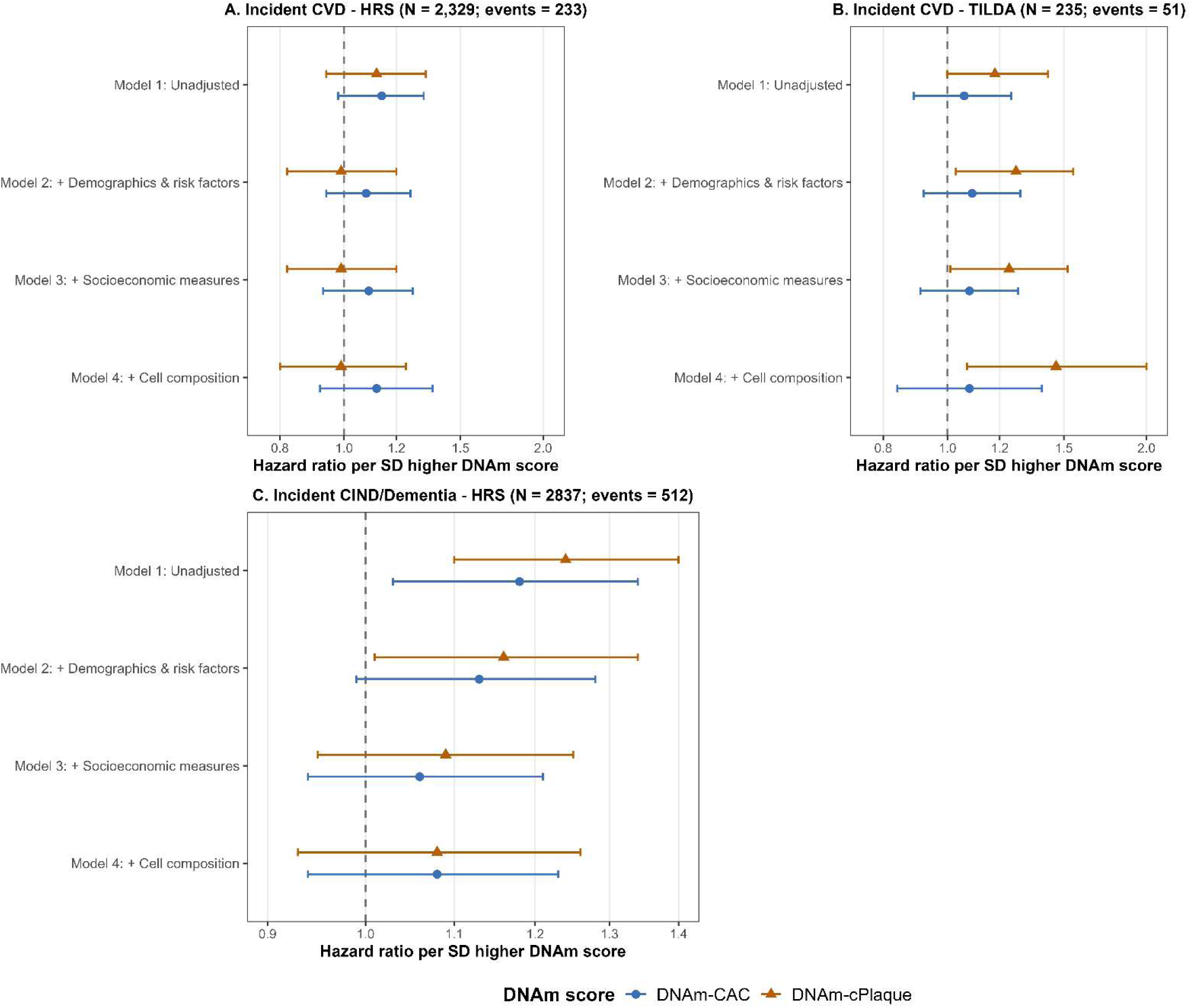
Associations of DNAm atherosclerosis scores with incident vascular outcomes in the HRS and TILDA Panels. **A** and **B** show incident cardiovascular disease in HRS and TILDA, respectively, and **Panel C** shows incident cognitive impairment, not dementia (CIND), or dementia in HRS. Effect estimates and 95% CIs are shown for DNAm-CAC and DNAm-cPlaque across sequential adjustment models. Incident cardiovascular disease was modeled using Cox proportional hazards models in HRS and interval-censored accelerated failure time models in TILDA (corresponding estimates are provided in **Table S3** and **Table S4**).

In TILDA, DNAm-CAC was not associated with incident CVD. In contrast, each SD increase in DNAm-cPlaque was associated with approximately 24% higher risk of incident CVD after adjustment for demographic and clinical risk factors, with effect estimates increasing to approximately 46% higher risk after additional adjustment for socioeconomic characteristics (Model 3). After further adjustment for leukocyte composition, this association was attenuated but remained directionally consistent with approximately 18% higher CVD risk. In sensitivity analyses using Cox models, results were largely consistent with the primary analyses. DNAm-cPlaque remained associated with incident CVD in fully adjusted models, whereas DNAm-CAC showed a marginal association **(eTable 5)**.

## DISCUSSION

In this cross-cohort analysis of older adults from the United States and Ireland, DNAm signatures of atherosclerosis were associated with multiple vascular-related outcomes, including CVD, cognitive impairment, and kidney function. The clearest longitudinal signal was observed for DNAm-cPlaque and incident cardiovascular disease, while cross-sectional associations with kidney function were evident for both DNAm-CAC and DNAm-cPlaque in TILDA. Across models, associations were strongest in minimally adjusted analyses and were attenuated after accounting for clinical risk factors and socioeconomic indicators. DNAm atherosclerosis scores were associated with vascular risk factors and socioeconomic characteristics in both cohorts, suggesting that these signatures may reflect accumulated vascular risk exposure and social patterning. Despite differences in cohort structure, outcome definitions, and sample size, associations were largely consistent in direction. Together, these findings suggest that DNAm signatures of atherosclerosis may capture molecular changes related to systemic atherosclerosis.

Several studies have examined associations between epigenetic biomarkers and cardiovascular, cognitive, and aging-related outcomes. One example of these biomarkers is epigenetic clocks which aggregate DNAm signals into composite measures of biological aging.^36–39^ DNAm-based scores trained on cardiovascular disease or cardiovascular health have also demonstrated modest improvements in risk prediction when added to traditional risk factor models.^40,41^ In this context, our study extends prior work by evaluating DNAm signatures derived from atherosclerosis phenotypes rather than clinical outcomes. Although these scores were not trained on the specific outcomes examined here, their associations with multiple vascular-related outcomes across cohorts suggest that they may reflect an underlying systemic atherosclerotic disease burden. Further research is needed to determine whether these DNAm signatures contribute to risk stratification and how they relate to established clinical and socioeconomic risk factors.

Atherosclerosis is a complex life-course process shaped by the cumulative influence of behavioral, environmental, and social risk factors that may not be fully captured by traditional clinical measures. DNAm may provide insight into this process because epigenetic variation reflects both genetic influences and biological responses to long-term exposures, including socioeconomic disadvantage and health-related behaviors.^42–49^ In our study, DNAm atherosclerosis scores were associated with established vascular risk factors, and effect estimates for vascular-related outcomes were substantially attenuated after adjustment for clinical and socioeconomic characteristics. This pattern supports the interpretation that DNAm signatures capture molecular correlates of cumulative vascular risk exposure rather than acting as independent causal drivers of disease. Peripheral blood represents a relevant tissue for studying these processes because circulating leukocytes participate directly in inflammatory and oxidative pathways central to atherosclerosis and are responsive to environmental and psychosocial stressors across the life course.^50,51^ Further work is needed to clarify biological pathways linking epigenetic processes with vascular risk exposures and disease progression across the life span.

Atherosclerosis at different vascular sites reflects varying biological aspects of atherogenesis, with each site demonstrating both shared and distinct relationships with clinical disease.^52–58^ Prior studies have shown that coronary artery calcium burden more strongly predicts coronary heart disease and composite cardiovascular events than carotid plaque measures, whereas predictive performance is more comparable for cerebrovascular outcomes. ^59,60^ In addition, atherosclerotic burden at vascular sites outside the coronaries has been linked to higher risk of dementia.^8,61,62^ These findings support the concept that markers of atherosclerosis at different vascular sites capture distinct dimensions of vascular disease risk. Consistent with this, DNAm-CAC in our study was more broadly related to vascular risk factors and cross-sectional indicators of vascular disease burden, whereas DNAm-cPlaque showed more selective associations across outcomes, including the most consistent longitudinal association with incident cardiovascular disease. Together, these patterns suggest that the two DNAm scores may capture overlapping but not identical dimensions of systemic vascular disease processes.

Associations between DNAm atherosclerosis scores and vascular-related outcomes varied across the HRS and TILDA cohorts, likely reflecting differences in cohort structure, outcome definitions, age distribution, and social and clinical risk factor profiles. For example, social patterning and associations with cognitive outcomes were more pronounced in HRS, whereas associations with kidney function and prospective cardiovascular disease risk, particularly for DNAm-cPlaque, were more evident in TILDA. Differences in confounding structures and measurement characteristics, including leukocyte composition adjustment in TILDA, may have contributed to variation in effect estimates. Differences in cardiometabolic risk profiles across cohorts may also partly account for heterogeneity in associations; for example, diabetes was more prevalent in HRS, whereas hypertension was more common in TILDA. Differences in sample size and number of incident events may also have contributed to variation in statistical significance across cohorts. Prior studies of epigenetic aging measures found more consistent associations across these two cohorts, though some of the outcome associations were reported for minimally adjusted models only.^63,64^ Importantly, the use of two population-based cohorts with longitudinal follow-up represents a major strength of this study, allowing evaluation of DNAm atherosclerosis signatures across different contexts. The availability of rich covariate data enabled careful assessment of attenuation patterns after adjustment for clinical and socioeconomic risk factors. These design features support the interpretation that DNAm signatures of atherosclerosis may capture aspects of cumulative vascular risk exposure that manifest differently across populations and outcome definitions.

This study has several limitations. First, DNAm atherosclerosis scores were derived using external weights from a cross-sectional epigenome-wide association study of atherosclerosis conducted in a disease-free cohort, which may limit generalizability to broader populations. That study evaluated DNAm in a single leukocyte cell type and had a modest sample size, potentially constraining the precision and biological scope of the derived scores. Second, cardiovascular disease was based on self-reported physician diagnoses, which may introduce misclassification despite prior evidence supporting reasonable validity for broadly defined cardiovascular phenotypes.^65,66^ Some associations were cross-sectional, limiting the ability to exclude reverse causation or subclinical disease at baseline. Although longitudinal analyses were conducted, the number of incident events and duration of follow-up were modest, which may have reduced statistical power. Residual confounding by unmeasured behavioral, environmental, or social factors cannot be ruled out and selective attrition over time may have influenced observed associations. Studies incorporating more rigorously adjudicated outcomes, larger samples, repeated DNAm assessments, and extended follow-up are needed to clarify temporal relationships, understand trajectories of epigenetic change across the lifespan, and evaluate whether DNAm signatures of atherosclerosis can inform mechanisms or preventive interventions.

In conclusion, DNAm signatures of atherosclerosis were associated with vascular risk profiles and multiple vascular-related outcomes across two population-based cohorts. Differences in associations across outcomes, models, and cohorts suggest that these signatures may capture distinct yet overlapping dimensions of vascular disease burden and progression. As epigenetic biomarkers are increasingly incorporated into population research, understanding how DNAm signatures reflect life-course exposures, systemic vascular aging processes, and modifiable risk pathways will be critical for evaluating their role in risk stratification and prevention of chronic disease.

## ACKNOWLEDGEMENTS

We thank the participants in the Health and Retirement Study and The Irish Longitudinal Study of Aging (TILDA) and the staff who catalogued their information.

## SOURCES OF FUNDING

This work was supported by the National Institute on Aging (U01AG009740, R01 AG060110, and R01 AG071071).

TILDA was supported by funding from Science Foundation Ireland (SFI) (SFI-19/US/3615), the Health Research Board (HRB) and an Irish Research Council (Advanced Laureate Award, IRCLA/2023/2029).

## DISCLOSURES

None

## SUPPLEMENTAL MATERIAL

Supplemental Methods

Tables S1 - S4

References

## DATA AVAILABILITY

Data from the Health and Retirement Study (HRS) are available to qualified researchers. Public-use data can be accessed through the HRS website, while DNA methylation data from the 2016 Venous Blood Study are available via the National Institute on Aging Genetics of Alzheimer’s Disease Data Storage Site (NIAGADS; https://www.niagads.org/qualified-access-data/) upon submission and approval of a Data Access Request and completion of required data use agreements.

Data from the The Irish Longitudinal Study on Ageing (TILDA) are available for non-commercial research use through application for data access and via a remote access platform, the TILDA VISTA Trusted Research Environment (https://tilda.tcd.ie/tilda-vista/).

## CODE AVAILABILITY

Codes will be made available upon reasonable request by the corresponding author.

## NONSTANDARD ABBREVIATIONS AND ACRONYMS

BMI: BODY MASS INDEX
CAC: CORONARY ARTERY CALCIFICATION
CIND: COGNITIVE IMPAIRMENT BUT NOT DEMENTIA
CPG: CYTOSINE-PHOSPHATE-GUANINE SITE
CVD: CARDIOVASCULAR DISEASE
DNAm: DNA METHYLATION
EWAS: EPIGENOME-WIDE ASSOCIATION STUDY
eGFR: ESTIMATED GLOMERULAR FILTRATION RATE
HRS: HEALTH AND RETIREMENT STUDY
MoCA: MONTREAL COGNITIVE ASSESSMENT
TILDA: THE IRISH LONGITUDINAL STUDY ON AGEING
VBS: VENOUS BLOOD STUDY

## Notes

### Competing Interest Statement

The authors have declared no competing interest.

### Author Declarations

The University of Michigan Institutional Review Board approved the collection and analyses of these data (HUM00061128, HUM00056464, and HUM00192059). Ethical approval for TILDA was granted by the Faculty of Health Sciences Research Ethics Committee in Trinity College Dublin.

